# Associations between the mid-adolescent external exposome and proteomic biomarkers of mental health

**DOI:** 10.1101/2024.12.18.24319208

**Authors:** Zhiyang Wang, Gabin Drouard, Aleksei Afonin, Núria Botella, Carmen Peuters, Aino-Kaisa Piironen, Alyce. M. Whipp, Boris Cheval, Libor Šulc, Marja Heinonen-Guzejev, Maria Foraster, John Gulliver, Jenny Selander, Payam Dadvand, Jordi Júlvez, Irene van Kamp, Katja M. Kanninen, Jaakko Kaprio, Equal-Life Scientific Team

## Abstract

The exposome, encompassing all external and internal environmental factors, provides a comprehensive understanding of the complex etiology of mental health. This study investigated the relationship between the external exposome during mid-adolescence and plasma proteomic biomarkers of mental health in 935 participants from the FinnTwin12 (Finland) and WALNUTs (Spain) cohorts. The mid-adolescent external exposome included 60 exposures. Biomarkers included 26 plasma proteins (mean age 22) for FinnTwin12 and 31 (mean age 13) for WALNUTs. The exposome-wide proteome-wide analysis revealed seven exposures (related to lifestyle, indoor environmental quality, natural environment, and family environment) linked to six proteins. Greenspace-related exposures were associated with proteins in both cohorts. However, these exposures showed no direct association with concurrent measures: the psychopathology factor or depressive symptoms. Twin analyses indicated genetic influences in some covariations between exposures and proteins. These findings provide evidence for mid-adolescents about the association between external environments and proteomic biomarkers of mental health.

## Main

Mental health conditions have emerged as a significant contributor to the global burden of disease^1^. Approximately one in eight people worldwide suffers from a mental health condition^1^, and the prevalence in adolescents is slightly higher (approx. one in seven)^2^. During adolescence, major physiological changes in the nervous and endocrine systems facilitate the development of coping strategies, interpersonal skills, and individual risk perception^3^. However, suboptimal timing or deviating magnitude of these changes can increase the risk of mental health conditions, making adolescence a vulnerable period^4,5^. In view of the complexity and overlap of mental health conditions, a general risk factor, known as the psychopathology factor (p-factor), was introduced for nonspecific mental health conditions by Caspi et al.^6,7^. A broad range of items measuring psychiatric symptoms was hereby aggregated into various distinct diagnoses, covering two overarching domains: Externalizing and Internalizing, and finally into the general latent dimension^6^. The p-factor unites all mental health conditions and has a neurobiological root^6^.

Besides being self-reported or reported by close relatives or professionals, an individual’s mental health status can also be reflected in molecules in blood, other body fluids, or tissues. Omics refers to the collective characterization and quantification of these molecules. Studies have identified molecules as biomarkers from different omics layers (e.g., proteomics) that are associated with mental health conditions ^8–10^. Biomarkers are further categorized based on their function. For example, diagnostic biomarkers help identifying patients with specific conditions and improve disease classification, which is crucial in mental health care due to its high heterogeneity, low inter-rater reliability, and common comorbidities^11^. Safety biomarkers, measured before and after exposure to environmental agents, can monitor adverse events, toxicity, and progression^11^. Overall, biomarker research is able to advance personalized psychiatry that develops preventive strategies, improves diagnosis and treatment, and understands the underlying mechanisms of mental health conditions.

Previous literature has underscored the role of environmental factors in adolescent mental health^12^. Departing from the traditional “one exposure, one disease” model, the exposome framework has been adopted, referring to the totality of environmental factors from a variety of external (not only the physical environment but also the socioeconomic context) and internal exposures individuals ever experienced^13^. Internal exposures are biomarkers of effect proven to be associated with external exposures. For example, cortisol is recognized as a biomarker for mental and psychiatric disorders, and some chemical exposures, such as pesticides, have been shown to relate with cortisol levels^14,15^. Thus, cortisol can be considered as an internal exposure. Under the exposome framework, various external exposures have been identified to associate with mental health including in the period of adolescence^16–20^. Although some studies have explore the relationship between the external exposome and multiomic profile^21–25^, research linking the external exposome, internal exposome/biomarkers, and mental health conditions remains relatively limited. Such research is crucial for understanding the complex biological networks through which environmental exposures connect to physiological changes.

When discussing the relationship between the environment and mental health, genetics should be considered, as many mental health conditions show heritability to different degrees^26^. The evaluation of interplay between genetic and environmental factors could help to elucidate individual differences in vulnerability and resilience to environmental stressors in relation with mental health. Research using gene-encoded proteins has further explored this interplay and revealed the potential of mental health biomarkers in such investigation^27^. Our previous research also highlighted a notable genetic contribution to the covariance between the exposome and depressive symptoms in adolescents and young adults^16^.

The main aim of this study was to evaluate the associations between the mid-adolescent external exposome and plasma proteomic biomarkers of mental health. Two types of exposome-wide proteome-wide analyses were used: multivariate Bayesian model with shrinkage priors (MBSP) for the one-to-multiple relationship and exposome-wide association study (ExWAS) for each protein for the one-to-one relationship. Second, we aimed to examine to what extent proteins explain the association between exposures and mental health and to investigate the genetic influences underlying significant exposure—protein associations using twin design. The analysis process is outlined in Figure 1.

**Fig. 1:**
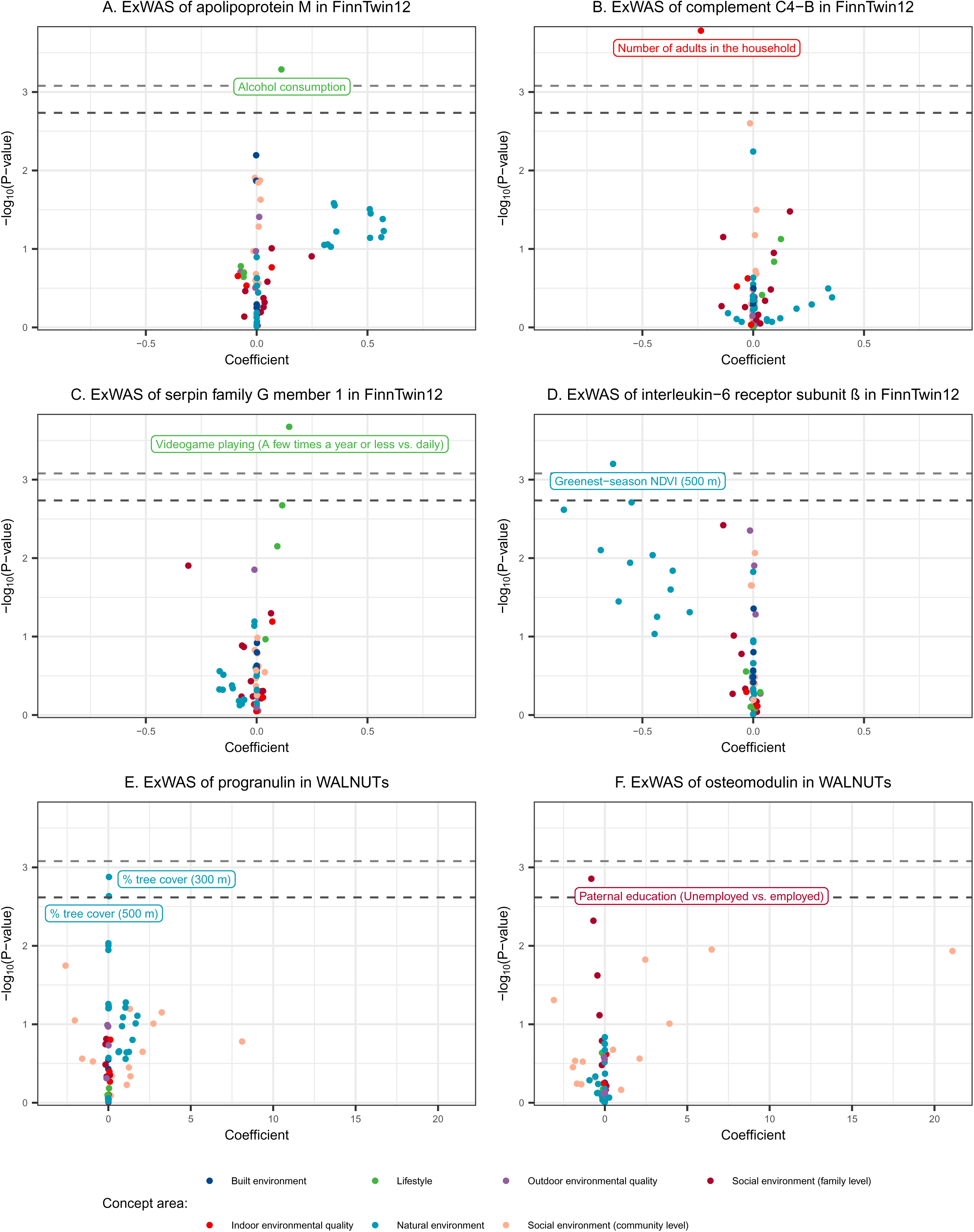
Flowchart of the study pipeline. Flowchart of the study demonstrating the path from the selection of external exposome and proteome to exposome-wide proteome-wide analysis and ending with twin analysis and pathway exploration toward mid-adolescent external mental health.

## Results

### Description of participants and materials

Participants included 748 individuals from FinnTwin12 (40.6% female) and 187 from WALNUTs (55.6% female) cohorts, with mean ages of 22.3 years (SD: 0.6) and 13.3 years (SD: 1.0), respectively, at plasma sampling for proteome assessment. Given that the included participants were a subsample of each cohort, Supplemental Table 1 presents the comparison of the distribution of sex and parental education level between the included and baseline-available samples.

The mid-adolescent external exposome set consisted of 60 exposures categorized into seven concept areas (Supplemental Table 2): built environment (5), social environment (community level) (14), social environment (family level) (6), indoor environmental quality (3), lifestyle (3), natural environment (25), and outdoor environmental quality (4).

Biomarkers for mental health (Supplemental Table 3) included 26 proteins in FinnTwin12 and 31 in WALNUTs, of which 25 were common for both cohorts. These protein biomarkers were selected with strong biological plausibility to mental health, and the detailed selection process is presented in the Method section.

### One-to-multiple relationships between exposures and proteins

Based on the MBSP^28^ adjusting for age at plasma sampling for proteome assessment and sex, we did not identify any exposure being associated with proteins in either cohort (by 95% posterior credible intervals). After further adjusting for body mass index (BMI), still, no significant associations were found in either cohort.

### One-to-one relationships between exposures and proteins

After Bonferroni correction by the number of effective test for multiple testing, the significant threshold for the P value was 1.87×10^-3^ in FinnTwin12 and 2.42×10^-3^ in WALNUTs. The overall results of ExWASes^16^ to each protein biomarkers are presented in Supplemental Tables 4 (FinnTwin12) and 5 (WALNUTs). Volcano plots in Figure 2 present those that have significant associations. Age at plasma sampling and sex were adjusted for.

**Fig. 2:**
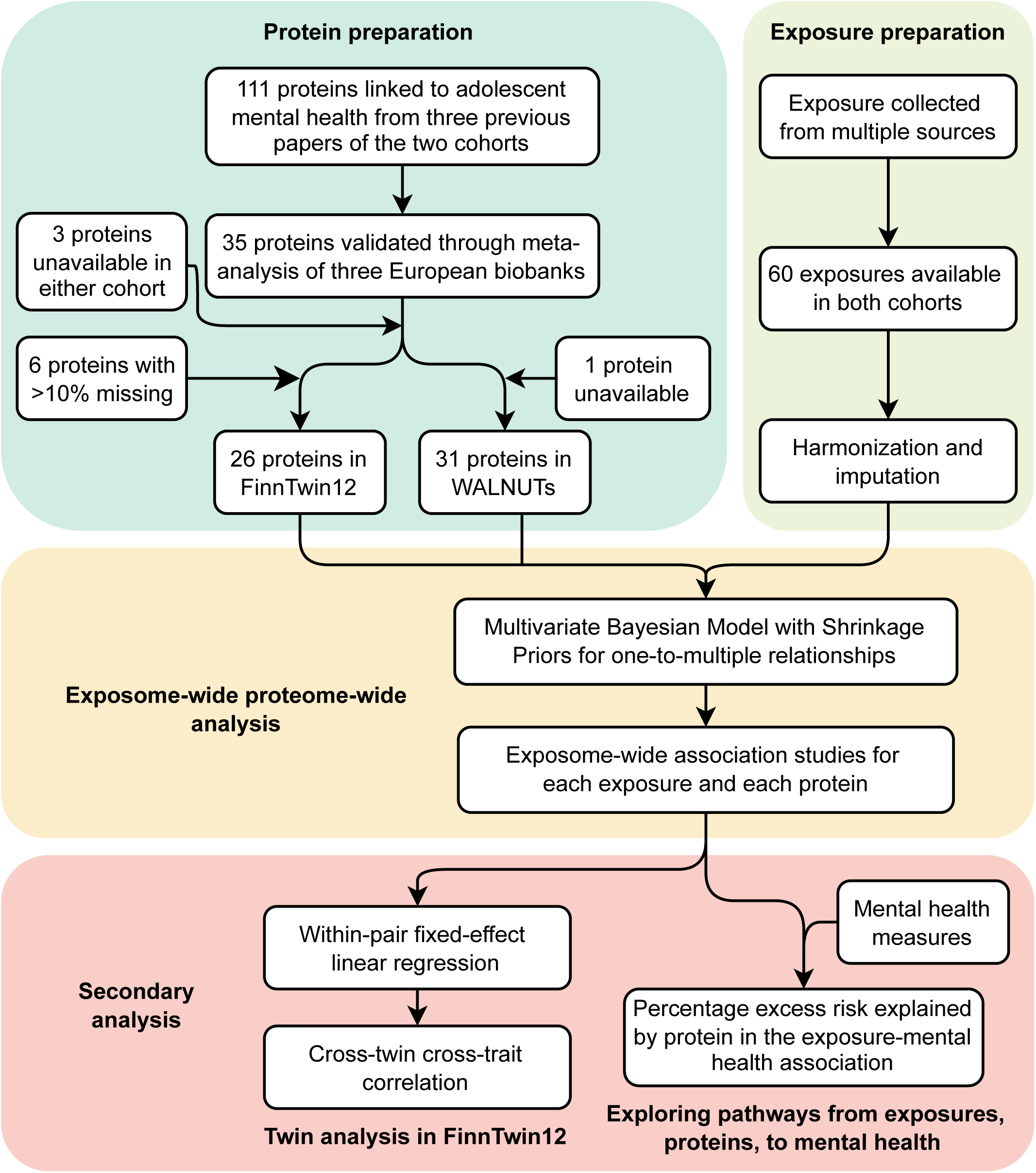
Volcano plots of exposome-wide association studies (ExWASes) for apolipoprotein M (A), complement C4-B (B), serpin family G member 1 (C), interleukin-6 receptor subunit β (D) in FinnTwin12 and progranulin (E) and osteomodulin (F) in WALNUTS. Only ExWASes with significant associations were plotted. The y-axis shows statistical significance as – log_10_(p-value) for the adjustment for multiple testing. Adjustments were made for sex and age at plasma sampling for proteome assessment. The significance thresholds for p-values were 1.84×10^-3^ for FinnTwin12 and 2.42×10^-3^ for WALNUTs (the dark gray dash line). The light gray dash line indicated the significance threshold by a restricted Bonferroni correction (p-value <8.33×10^-4^).

In FinnTwin12, four exposures in three concept areas (two in lifestyle, one in indoor environmental quality, and one in natural environment) were significantly associated with four different proteins. Participants who reported consuming alcohol at age 14 had higher levels of apolipoprotein M compared to non-consumers (Beta: 0.11, 95% confidence interval (CI) 0.05, 0.17) (Figure 2A). A higher number of adults in the household (household size) when participants were 11–12 years old was associated with lower levels of complement C4-B (Beta: -0.34, 95% CI: -0.36, -0.11) (Figure 2B). Participants who played videogames daily at age 14 had higher levels of serpin family G member 1 compared to those who played a few times a year or less (Beta: 0.15, 95% CI: 0.07, 0.22) (Figure 2C). Moreover, the greenest-season Normalized Difference Vegetation Index (NDVI) within a 500 m buffer was negatively associated with interleukin-6 receptor subunit β (Beta: -0.63, 95% CI: -0.99, -0.27) (Figure 2D).

In WALNUTs, three exposures in two concept areas (two in natural environment, one in social environment at the family level) were significantly associated with two proteins. The percentages of the area covered by trees within 300 (Beta: 0.04, 95% CI: 0.01, 0.06) and 500 (Beta: 0.03, 95% CI: 0.01, 0.05) m buffers were positively associated with progranulin (Figure 2E). Participants whose fathers were unemployed had lower levels of osteomodulin (Beta: -0.82, 95% CI: -1.32, -0.32) compared to those whose fathers were employed (Figure 2F).

The statistical power of significant associations were all 1.00 in FinnTwin12 and ranged from 0.15 to 1.00 in WALNUTs.

### Secondary analysis

First, we assessed to what extent the protein explains the association between exposure and mental health (measured at the same time as plasma sampling) based on the significant ExWAS results. A total of 733 out of 748 participants in FinnTwin12 and all included participants in WALNUTs provided valid mental health measures of depressive symptoms based on the General Behavior Inventory (GBI) and the p-factor based on the Strengths and Difficulties Questionnaire (SDQ), respectively. In both FinnTwin12 and WALNUTS, none of these exposures was significantly associated with either depressive symptoms or the p-factor, and the protein explained only a very small portion of weak associations between exposures and mental health measures (Extended Figure 1).

Second, we assessed the genetic contribution by twin analyses. In the within-pair fixed-effect regression model, only the association between videogame playing and serpin family G member 1 remained significant (Beta: 0.19, 95% CI: 0.05, 0.34) (Extended Figure 2A, Supplemental Table 6). After keeping only full pairs, the cross-twin cross-trait (CTCT) correlation coefficient between videogame playing and serpin family G member 1 in monozygotic (MZ) twins was almost 5 times higher than in dizygotic (DZ) twins. The CTCT correlation coefficient between alcohol consumption and apolipoprotein M was nearly two times higher in MZ pairs than in DZ pairs (Extended Figure 2B). Supplemental Note 1 illustrated more on results and methodology.

### Sensitivity analysis

The sensitivity ExWASes that further adjusted for BMI were presented in Supplemental Note 2 and Extended Figure 3. Four exposures in the concept area of the social environment (community level) were significantly associated with desmoglein-2 in FinnTwin12, while one exposure in the same concept area was significantly associated with fibulin-1 in WALNUTs.

The sensitivity ExWASes based on the young adulthood external exposome (50 exposures) were shown in Supplemental Note 2 and Extended Figure 4. Alcohol consumption frequency reported in young adulthood was significantly associated with six proteins including apolipoprotein M. Videogame playing was associated with transferrin. Five exposures in the concept area of the natural environment were associated with lysosome-associated membrane glycoprotein 1. The annual mean concentration of black carbon was associated with IgGFc-binding protein.

Due to the circannual dramatic difference in daylight in Finland, we assessed the interaction effect of the season when participants responded to the questionnaire on the association between videogame playing and serpin family G member 1. However, no significant interaction was found (Supplemental Note 2 and Supplemental Table 7).

## Discussion

This study, involving nearly a thousand participants from Finland and Spain, systematically explored the relationship between various external environmental exposures during mid-adolescence and plasma proteomic biomarkers of mental health. Seven exposures in the concept areas of lifestyle, indoor environmental quality, natural environment, and social environment (family level) were significantly associated with six different proteins. Notably, greenspace-related exposures consistently showed associations with proteins in both countries. None of these seven exposures were further significantly associated with mental health conditions. Additionally, twin analysis revealed some genetic influence on the relationship between exposures and proteins, particularly between videogame playing and serpin family G member 1.

This study confirms previous findings regarding associations between external exposures and plasma proteins. First, apolipoprotein M, a lipocalin protein linked to high-density lipoprotein (HDL), was found to be associated with alcohol consumption in mid-adolescence, as well as with consumption frequency in young adulthood. Consumption frequency in young adulthood was moreover correlated with more proteomic biomarkers. A Danish case-control study suggested it significantly dysregulated the fibrosis stage among adult patients with alcohol-related liver disease compared to healthy controls^29^. A systematic review highlighted a positive correlation between alcohol consumption and all HDL subfractions^30^. A US cohort study further demonstrated the interaction effect of alcohol use on the relationship between platelet–HDL ratio and depressive symptoms in adults^31^. Early onset of alcohol use and misuse is also a predictor of mental health conditions in adulthood^32,33^. Osteomodulin was correlated with parental education level. In FinnGen, the gene coding for osteomodulin, OMD, shows a strongest association with height, which height is an important determinant of higher educational level, though the mechanism is not fully known^34,35^.

Next, we found significant associations between exposures and two proteins involved in the complement system, potentially contributing to mental health conditions through immune dysfunction. Complement C4−B, an immune effector, was negatively associated with household size. Infants in larger households were observed to have higher levels of regulatory cells and C-reactive protein during their first year, contributing to nonspecific immune responses^36^. Our findings suggest that smaller household sizes are associated with higher C4-B levels, which may alleviate mental health conditions, while contradicting the WHO reporting that crowded living conditions exacerbated mental health conditions^37,38^. The difference could be explained by unmeasured factors such as room availability. Household size is not equivalent to crowding, and the beneficial effect may be rooted in the close family atmosphere. Besides, Complement C4 genes are strongly associated with schizophrenia^39^ and predict suicide among schizophrenia patients^40^, suggesting them as a promising biomarker for psychiatric diseases^41,42^. Serpin family G member 1 was found to be associated with videogame playing in mid-adolescence. This protein engages in the regulation of the complement cascade, where elevated blood pressure activates the complement system and produces complement factors^43^. A cross-sectional study of 282 overweight or obese Canadian adolescents found that videogame playing was associated with higher blood pressure^44^. This rise in blood pressure may serve as a critical link between videogame playing, serpin family G member 1, and mental health conditions^45^. In addition, the serpin proteins are regarded as effective regulators of immune response and involved in neuronal signaling.

In the present study, exposures related to greenspaces were associated with protein levels in both countries, implying a universal connection. A US cross-sectional study found that distance-weighted vegetated land cover correlated with serum interleukin-8, which belongs to the same family as interleukin-6 receptor subunit β (finding in FinnTwin12)^46^. Both interleukin-6 receptor subunit β and progranulin, identified in our study, play roles in the immune system, indicating a potential biological mechanism behind this association. Although a proteome study about NDVI and coronary heart disease did not include overlapping proteins with our results, some proteins they identified were related to immune response and inflammation as well^47^. The microbial input from greenspaces helps the immune system to distinguish between harmful and beneficial antigens, which may affect mental health through reducing systemic inflammation^48^. However, combined with previous protein—mental health research, in brief, increased greenspaces were linked to worse mental health in Finland, while the opposite was observed in Spain^38,49^. This discrepancy may be due to differences in greenspaces’ coverage, type, or quality, as well as other contextual differences^50,51^. A recent systematic review indicated that higher quality greenspaces are generally associated with better mental health outcomes^52^. Better measurements to characterize greenspaces are desired. Our findings endorse the importance of greenspace in urban planning and encourage allowing children and adolescents to spend time in green environments.

None of the significant exposures were further associated with mental health conditions. Biomarkers, as early indicators, may reflect subclinical changes that have not yet progressed to noticeable symptoms or a diagnosable condition. A global meta-analysis found that the peak onset for mental health conditions such as primary psychotic states and mood disorders occurs around age 20^53^. This suggests that symptoms or conditions may develop later than our measurement time, especially during mid-adolescence in WALNUTs. The very small PERE value further suggested that these exposures were pleiotropic, which might be linked to mental health via multiple proteins or other molecules. For example, research using phenome-wide association studies and Mendelian randomization has shown that polymorphisms in genes encoding alcohol-metabolizing enzymes is correlated with various health domains, including mental health, implying the involvement of multiple proteins^54^. Our current proteomic analysis is specific to mental health, and the pathway was through single selected proteins, therefore future studies should expand the proteomic scope and integrate other omics layers to construct a broader picture.

Genetic influences are notable in the associations between exposure and proteins. After adjusting for genetic effects, videogame playing remained associated with serpin family G member 1, while their CTCT correlation coefficient was much higher in MZ than in DZ. Although not all genetic and shared environmental effects were controlled, this finding indicates that environmental factors coexist with genetic influences. Univariate twin modeling has indicated the genetic variance in the proteome^55,56^, alcohol consumption^57,58^, and videogame playing^59^. This finding highlights that genes and environment are not an either-or opposition but intertwined. In twin studies, the genetic effect is referred to as the heritable effect passed down through generations within families. Considering family as part of the exposome, it emphasizes the inseparability of the gene and environment and their intricate connection with phenotype^16^. Twin studies applying proteomics further elucidate the genetic nurture hypothesis, which explains how parental environmental influences shape offspring outcomes, providing a biological rationale^60^.

Some limitations were noted. First, the relatively small sample size could have limited the statistical power with higher risks of type I error and likelihood of null findings. Although most of the significant associations from ExWASes have adequate statistical power, the weak power caused by small effect size in WALNUTs’ findings highlights the concern of the ubiquity of the exposome, wherein numerous exposures may play small yet significant roles that we may have overlooked. Collaboration with more cohorts is encouraged. Second, calculating the PERE by protein represents an exploratory approach aimed at linking exposures, proteins, and mental health outcomes but lacks causal inference capability. The interpretation should be cautious. High-dimensional mediation analysis techniques are encouraged for complexities of omics data. Third, our analyses did not account for co-exposure scenarios, which may raise concerns on confounding, interaction, or other issues. Future research integrating various exposures into a single analytical framework could assess the mixture, joint, or combined effects of these exposures to provide a more comprehensive understanding of the effect of the environmental totality. Fourth, the exposome set included in our study comprised only those exposures available in both cohorts, which may not encompass all potential exposures, and a larger inclusion of exposure can enhance the systemic investiagtion of the environmental influence in the future.

## Conclusion

Seven external exposures during mid-adolescence related to lifestyle, indoor environmental quality, natural environment, and social environment at the family level were associated with six proteomic biomarkers of mental health. Effects from greenspaces were observed in both Finland and Spain. Although these biomarkers have been proven to be related to mental health with strong biological plausibility, these exposures were not directly associated with depressive symptoms or the p-factor measured simultaneously with plasma sampling. Twin analysis highlighted the interplay between genetic and environmental factors in shaping these associations. These findings offer more comprehensive insights into how environmental factors are linked to proteomic biomarkers of mental health, even before the onset of mental health conditions.

## Methods

### Participants

This study was based on harmonized data from two cohorts, FinnTwin12 and WALNUTs. FinnTwin12 is a population-based prospective cohort of all Finnish twins born between 1983 and 1987, consisting of one baseline and four follow-ups currently. When twins reached age 14, 1035 families with 2070 twins were invited to an intensive sub-study with psychiatric interviews, biological sampling, and additional questionnaires, and 1854 twins participated. They were then invited to participate again as young adults (mean age 21.9), with 73% completed. The subsample was mostly randomly selected from the full sample, while enriched with twins at elevated familial risk for alcoholism. The updated overview of FinnTwin12 has been published^61^. The WALNUTs cohort consists of 771 adolescents who participated in a multi-school, parallel, controlled, 6-month superiority randomized clinical trial in Barcelona, Spain, with baseline measurement between April 2016 and June 2017^62^. Inclusion criteria were attending regular secondary education (age 11-16) in Barcelona, Spain. Exclusion criteria were regular consumption of omega-3 PUFAs supplements, daily consumption of walnuts, lactose intolerance, or allergy to walnuts, gluten, cereals, dried fruits, peanuts, soy, sesame, or sulfites.

The inclusion criteria of this study were: 1) available plasma sample and untargeted analysis of plasma proteome, 2) data on residential history, and 3) data on mental health and exposure from at least one cohort questionnaire. A total of 748 participants from FinnTwin12 and 187 participants from WALNUTs were included. Comparing the included and full baseline sample, the distribution of sex were similar in both cohort (Supplemental Table 1). A higher proportion of females in FinnTwin12 and males in WALNUTs were included. In FinnTwin12, this may be because we non-randomly included twins at elevated familial risk for alcoholism in the intensive sub-study^63^. In WALNUTs, we only included participants with available proteome assay^62^.

The ethics committee of the Department of Public Health of the University of Helsinki (Helsinki, Finland) and the Institutional Review Board of Indiana University (Bloomington, Indiana, USA) approved the FinnTwin12 study protocol from the start of the cohort. The ethical approval of the ethics committee of the Helsinki University Central Hospital District (Helsinki, Finland) (HUS) is the most recent and covers the most recent data collection (wave 4) (HUS/2226/2021). The HUS reviews the study annually, and 2024’s statement is number 14/2024, dated May 22^nd^, 2024. All participants and their parents/legal guardians gave informed written consent to participate in the study. The WALNUTs study was approved by the CEIC Parc Salut Mar Clinical Research Ethics Committee (approval number: 2015/6026). The trial was registered at ClinicalTrials.gov (NCT02590848) on 29/10/2015. Written informed consent to participate in the original WALNUTs study was provided by the participants’ legal guardian/next of kin. The authors assert that all procedures contributing to this work comply with the ethical standards of the relevant national and institutional committees on human experimentation and with the Helsinki Declaration of 1975, as revised in 2008.

### External exposome

A total of 60 external exposures were curated from both cohorts and classified into seven categories. The description and data source can be found in Supplemental Table 2. We only included exposures with less than 20% missing data in a practical way to balance between losing too much information and maintaining the integrity of the dataset. We selected the exposure period closest to the mid-adolescence (age 14 in FinnTwin12 and age 11-16 in WALNUTs). Exposures in the concept areas of social environment (family level), indoor environmental quality, and lifestyle were collected via questionnaires in each cohort. Social environment (community level) exposures were derived from national statistics agencies at the postal code level in Finland and at the census section level in Barcelona. Exposures related to the built environment, natural environment, and outdoor environmental quality were obtained from open-source platforms such as the Copernicus program or were calculated based on land use regression model, described elsewhere^13,16,64^.

For example, NDVI quantified vegetation greenness and characterize the vegetation density, derived from U.S. Geological Survey Landsat data via Google Earth Engine^65^. These exposures were at the geocode level. Residential history from the national registry up to 2020 in FinnTwin12 and the current and one-year previous residential address inquired from parents during baseline in WALNUTs was used to integrate exposures into the cohorts, if these exposures were not collected via questionnaires. We imputed missing values with the median in each cohort.

### Proteomic biomarkers of mental health

We used plasma samples obtained in young adulthood (mean age: 22) in FinnTwin12 and at the baseline (age 11-16) in WALNUTs to profile the proteome. The detailed procedure for sample handling is presented in previous studies^49,66^. For this study the raw MS/MS files were re-analysed with Spectronaut software (Biognosys; version 18.0.2). The direct DIA approach was used to identify proteins. The main data analysis parameters in Spectronaut were: (i) Enzyme: Trypsin/P; (ii) up to 2 missed cleavages; (iii) Fixed modifications: Carbamidomethyl (cysteine); (iv) Variable modifications: Acetyl (protein N-terminus) and oxidation (methionine); (v) Precursor FDR Cutoff: 0.01; (vi) Protein FDR Cutoff: 0.01; (vii) Quantification MS level: MS2; (viii) Quantification type: Area under the curve within integration boundaries for each targeted ion; (ix) Protein database: Homo sapiens Swiss-Prot reference proteome (Uniprot release 2022_01_07_HUMAN)^67^ and Universal Protein Contaminant database^68^. As both datasets were run in multiple batches, batch correction was performed using the proBatch package. The batches of FinnTwin12 and of WALNUTs have been described elsewhere^66,69^.

Then, based on three previous proteome studies on both cohorts, 111 proteins were identified that were associated with adolescent mental health^38,49,66^. The coding genes of these proteins were validated using meta-analyzed genetic association results from FinnGen, UK Biobank, and Estonia Biobanks (release 11) via FinnGen’s browser, ensuring biological plausibility with mental health and psychiatric outcome^34^. Validation was determined by significant (P-value < 5×10^-8^) associations of the protein’s coding gene with phenotypes falling into five categories: 1) comorbidities of neurological endpoints, 2) neurological endpoints, 3) diseases of the nervous system, 4) mental and behavioral disorders, and 5) psychiatric endpoints. Proteins identified by at least two studies were also considered validated. A total of 35 proteins met this criterion. After excluding proteins that were unavailable or had over 10% missing data, 26 and 31 proteins were included as biomarkers of mental health in FinnTwin12 and WALNUTs, respectively (Supplemental Table 3). For proteins with less than 10% missing data, they were imputed by the minimum observed value and log2-transformed^38^.

### Mental health measures

The p-factor was the main mental health outcome in this study, which summarizes occurrences of internalizing and externalizing symptoms and the propensity to develop any and all forms of common psychopathologies^6^. In WALNUTs, the p-factor was derived from the 10 internalizing and 10 externalizing problem items from the Spanish version of the Strengths and Difficulties Questionnaire (SDQ), rated on a 3-point Likert scale to assess emotional and behavioral difficulties^70^. We first calculated the mean scores for internalizing and externalizing problem items, allowing for up to four missing values per category (eight in total). Next, we added one to the mean score, applied a logarithmic transformation, and standardized it to z-scores, resulting in the p-factor.

However, there was no assessment of the p-factor in the FinnTwin12 study when participants were young adults. Instead, the short-version General Behavior Inventory (GBI) score, which assesses the occurrence of depressive symptoms, was used as a substitute for the p-factor^71^. The self-reported GBI consists of ten Likert-scale items, with total scores ranging from 0 to 30. Higher scores indicate a greater occurrence of depressive symptoms. The sum score was standardized too. Since depression is one key dimension of the p-factor, GBI serves as a suitable replacement in this context.

### Multivariate Bayesian model with shrinkage priors (MBSP)

The MBSP was employed to identify exposures affecting the majority of proteins (one-to-multiple relationship). It is a multivariate Bayesian technique with global–local shrinkage priors to generate a sparse model of predictors affecting multiple outcomes^72^. The model has a good performance and selects variables by examining the 95% posterior credible interval^28^. There were nine sets of different priors and tau inputted into models, and the Deviance Information Criterion was used to identify the model with the best goodness-of-fit^72^. Two covariates were added: age at plasma sampling and sex. MBSP were performed separately for the two cohorts. The MBSP package in the R environment (version 4.3.3) was used^28^.

### Exposome-wide association studies (ExWASes)

Generalized linear regression models with Gaussian distribution were fitted between each exposure and protein, as ExWASes, to explore one-to-one relationships. Bonferroni correction by the number of effective tests was used to adjust for multiple testing, accounting for correlation in exposures^73^. The models were adjusted for the same covariates as the MBSP. To adjust for the clustered nature of the data (twins in FinnTwin12, schools in WALNUTs), the robust standard error was calculated. ExWASes were performed separately for the two cohorts. We used the rexposome package in the R environment (version 4.3.3)^74^.

### Percentage excess risk explained (PERE)

To assess the extent to which proteins explained the association between exposure and mental health measures, PERE was calculated based on generalized linear regression. We performed two models with the following independent variables of 1) exposure only and 2) exposure and protein^75^. The PERE formula is:

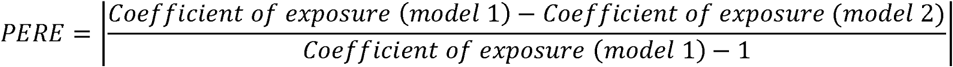

The PERE value could be interpreted as the involvement of proteins on the association between exposure and mental health measures, regardless of underlying mechanisms such as mediation and moderation^75^. PEREs were calculated based on the exposure—protein associations that were identified in ExWASes. The models were adjusted for age at plasma assessment and sex. Participants missing the mental health measures were excluded (7 in FinnTwin12). The standard error was robusted. The models were performed using Stata 18.0 (StataCorp).

### Twin analysis

The twin analysis was based on the significant exposure—protein associations identified in FinnTwin12. There were two parts: within-pair fixed-effect linear regression models (based on all included participants) and the cross-twin cross-trait (CTCT) correlations (based on the full twin pairs). The detailed methodology is presented in Supplemental Note 2.

### Sensitivity analysis

First, due to the significant influence of BMI on both the exposome^76^ and proteome^77^, we further added BMI at plasma sampling as another covariate in the MBSP and ExWAS. Participants without BMI information were excluded (N=19 in FinnTwin12), since BMI is the focus of sensitivity.

Second, given the gap between the timing of the exposome assessment (mid-adolescence) and plasma sampling (early adulthood) in FinnTwin12, we assessed 50 exposures to characterize the young adulthood exposome close to age 22, which were repeated measures of mid-adolescence exposures. Videogame playing was not inquired about in young adulthood and we replaced it with the videogame playing reported at age 17. We applied the young adulthood exposome to conduct ExWAS based on data from 768 participants.

## Data Availability

All data produced in the present study are available upon reasonable request to the authors.

## Acknowledgement

We would like to thank Dr. Stephanie Zeller, University of Helsinki, and Dr. Augusto Anguita-Ruiz, ISGlobal, for his support in the outcome-wide analysis.

## Author contribution

All authors conceived and conceptualized the research idea and developed the analysis plan. Z.W., M.F., J.G., J.J., and J.K. acquired the cohort or exposure data. A.A., A.P., and K.K. performed the proteome assessment. Z.W. led the statistical analysis and performed the analysis in FinnTwin12. N.B. and C.P. contributed to the statistical design and performed the analyses for WALNUTs. Z.W. drafted the original draft of the paper, and all authors reviewed the paper and provided critical comments. I.v.K., J.S., K.K., J.J., and J.K. gave critical supervision and guidance. All authors approved the paper.

## Data availability

The FinnTwin12 data are not publicly available due to the restrictions of informed consent. However, the FinnTwin12 data are available through the Institute for Molecular Medicine Finland (FIMM) Data Access Committee (DAC) for authorized researchers who have IRB/ethics approval and an institutionally approved study plan. To ensure the protection of privacy and compliance with national data protection legislation, a data use/transfer agreement is needed, the content and specific clauses of which will depend on the nature of the requested data. Requests will be addressed in a reasonable time frame (generally two to three weeks), and the primary mode of data access is by either personal visit or remote access to a secure server.

The WALNUTs data are not publicly available due to the restrictions of informed consent. The data contain personal information on children and, according to the ethical approval, they should be kept confidential. Data can be made available upon reasonable request to Jordi Júlvez for researchers who meet the criteria for access to confidential data. A data-use/transfer agreement is needed to ensure the protection of privacy and compliance with national data protection legislation, the content and specific clauses of which will depend on the nature of the requested data.

## Code availability

The main code for major analysis was uploaded to Github (https://github.com/doge73/exposome_proteome_mental/).

**Extended Fig. 1: Bar plots for protein-explained excess risk for exposure–mental health association based on the identified exposure–protein associations**

**a.** Percentage excess risk explained (PERE) by apolipoprotein M in the association between alcohol consumption and GBI score. **b.** PERE by complement C4-B in the association between the number of adults in the household and GBI score. **c.** PERE by serpin family G member 1 in the association between videogame playing and GBI score. **d.** PERE by interleukin-6 receptor subunit ß in the association between greenest-season NDVI (500m) and GBI score. **e.** PERE by progranulin in the association between % tree cover (300 m) and p-factor. **f.** PERE by progranulin in the association between % tree cover (500 m) and p-factor. **g.** PERE by osteomodulin in the association between paternal education and p-factor. The comparison was between participants playing video games every day and a few times a year or less (ref.) for the association between video game playing and GBI score and was between participants whose fathers received low and high (ref.) levels of education for the association between paternal education and p-factor.

**Extended Fig. 2: Twin analyses for the identified exposure–protein associations.**

**a.** Forest plot based on within-pair fixed-effect linear regression models. The comparison was between participants playing video games every day and a few times a year or less (ref.) for the association between videogame playing and serpin family G member 1. **b.** Histogram plot for cross-twin cross-trait correlation. Categorical exposure of videogame playing was regarded as a complete variable in the correlation calculation. Since the number of adults in the household remained constant between cotwins, no twin analysis was conducted to examine its association with complement C4-B.

**Extended Fig. 3: Volcano plots of sensitivity exposome-wide association studies (ExWASes) for apolipoprotein M (A), complement C4-B (B), serpin family G member 1 (C), interleukin-6 receptor subunit β (D), and desmoglein-2 (E) in FinnTwin12 and progranulin (F), osteomodulin (G), and fibulin-1(H) in WALNUTs**

Only ExWASes with significant associations were plotted. Adjustments were made for sex and age and BMI at plasma sampling for proteome assessment. The y-axis shows statistical significance as -log10(p-value) for the adjustment for multiple testing.

**Extended Fig. 4: Volcano plots of sensitivity exposome-wide association studies (ExWASes) for the young adulthood exposome for apolipoprotein M (A), transferrin (B), serpin family A member 6 (C), clusterin (D), lysosome-associated membrane glycoprotein 1 (E), fibulin-1 (F), serpin family A member 10 (G), and IgGFc-binding protein (H).**

Only ExWASes with significant associations were plotted. The young adulthood exposome were used instead. Adjustments were made for sex and age and at plasma sampling for proteome assessment. The y-axis shows statistical significance as -log10(p-value) for the adjustment for multiple testing.

