## Supplemental Note 1-2, Supplemental Table 1-7; Extended Figure 1-4 for "Associations between the mid-adolescent external exposome and proteomic biomarkers of mental health": prot_exp_suppl.docx

**Supplemental Note 1: Twin analysis in FinnTwin12: methodology and results.**

Methodology

First, within-pair fixed-effect linear regression models were employed, representing associations conditional on exposures shared within pairs and controlling for unobserved individual heterogeneity from genetics and shared environments^1^. It is a type of multilevel mixed model that removes any variation between cotwins as the fixed effects. The models were performed on all included participants. Age at plasma sample and sex were controlled for. The models were performed using Stata 18.0 (StataCorp).

Second, the cross-twin cross-trait (CTCT) correlations, stratified by zygosity, were calculated based on full twin pairs (N=325). The CTCT correlation showed the magnitude that exposure in twin 1 is correlated with the corresponding protein level in the cotwin^2^. It shows the extent to which individual differences in variables and covariation in these variables are attributed by genetic and environmental factors^2^. If the CTCT correlations were higher for monozygotic (MZ) twin pairs than for dizygotic (DZ) twin pairs, especially two-fold higher, it indicates a strong influence of genetic effects on the association between the exposure and protein^3^. We used the OpenMX package in the R environment (version 4.3.3)^4^.

If exposures were obtained from parent’s questionnaires, there was no twin analysis due to the invariability. Results involving enriched exposure based on residential address should be viewed with caution since most of cotwins lived together in mid-adolescence.

Result

Participants who reported consuming alcohol at age 14 had higher levels of apolipoprotein M compared to non-consumers (Beta: 0.09, 95% CI -0.03, 0.20) without strong evidence (P-value=0.14) (Extended Figure 2A). The CTCT correlation coefficients between alcohol consumption and apolipoprotein M were 0.09 in MZ and 0.05 in DZ pairs (Extended Figure 2B). Participants who played videogames daily at age 14 had significantly higher levels of serpin family G member 1 compared to those who played a few times a year or less (Beta: 0.19, 95% CI: 0.05, 0.34) (Extended Figure 2A). The CTCT correlation coefficients between videogame playing and serpin family G member 1 were 0.22 in MZ and 0.05 in DZ pairs (Extended Figure 2B). The greenest-season NDVI within a 500 m buffer was positively but insignificantly associated with interleukin-6 receptor subunit β (Beta: 2.39, 95% CI: -2.45 7.25) (Extended Figure 2A). Meanwhile the CTCT correlation coefficients were similar between MZ (0.20) and DZ pairs (0.20) (Extended Figure 2B).

**Supplemental Note 2: Sensitivity analysis**,

First, compared to main ExWASes, we additionally adjusted for BMI in ExWAS. The significance threshold in FinnTwin12 became 1.83×10^-3^, while there were no changes of the significance threshold in WALNUTs. In FinnTwin12 (sample size became 729), there were four exposures in the concept of the community social environment: percentage of community residents aged 30-39 (Beta: -0.02, 95% CI: -0.04, -0.01), 80-89 (Beta: 0.03, 95% CI: 0.01, 0.05), 0-9 (Beta: -0.02, 95% CI: -0.03, -0.01), and over 66 (Beta: 0.01, 95% CI: 0.00, 0.01) that were additionally associated with desmoglein-2 (Extended Figure 3E). Besides, participants who played videogames a few times a month at age 14 also had higher levels of serpin family G member 1 compared to those who played a few times or less (Beta: 0.13, 95% CI: 0.05, 0.20) (Extended Figure 3C). In WALNUTs, the percentage of community residents born abroad in the concept of the community social environment, was significantly associated with fibulin-1 (Beta: 2.16, 95% CI: 0.96, 3.37) (Extended Figure 3H). The effect sizes of other significant exposures were similar between the main and sensitivity analyses.

Second, we preformed ExWASes based on the young adulthood external exposome in FinnTwin12, The significance threshold in FinnTwin12 became 2.77×10-3. Compared to participants who reported consuming alcohol once a week in young adulthood, those who reported consuming alcohol 2-4 times a year or less, or even never in young adulthood had lower levels of apolipoprotein M (Beta: -0.22, 95% CI: -0.33, -0.11) (Extended Figure 4A), serpin family A member 6 (Beta: -0.26, 95% CI: -0.40, -0.12) (Extended Figure 4C), lysosome-associated membrane glycoprotein 1 (Beta: -0.11, 95% CI: -0.18, -0.05) (Extended Figure 4E), and serpin family A member 10 (Beta: -0.22, 95% CI: -0.33, -0.11) (Extended Figure 4G) and had higher levels of fibulin-1 (Beta: 0.22, 95% CI: 0.09, 0.35) (Extended Figure 4F). And, participants who reported consuming alcohol about once in one or two months have lower levels of clusterin (Beta: -0.10, 95% CI: -0.15, -0.04) (Extended Figure 4D). Compared to participants who reported playing videogame once in six months or less at age 17, those who played videogame once a few times per week (Beta: -0.16, 95% CI: -0.24, -0.08) and daily (Beta: -0.14, 95% CI: -0.23, -0.05) had lower levels of transferrin (Extended Figure 4B). The elevation within 100 (Beta: <0.01, 95% CI: <0.01, <0.01), 300 (Beta: <0.01, 95% CI: <0.01, <0.01), 500 (Beta: <0.01, 95% CI: <0.01, <0.01) m buffers were positively associated with lysosome-associated membrane glycoprotein 1 but in very small effect sizes (Extended Figure 4E). In addition, the percentages of the area covered by trees within 100 (Beta: <0.01, 95% CI: <0.01, <0.01) and 500 (Beta: <0.01, 95% CI: <0.01, 0.01) m buffers were also positively associated with lysosome-associated membrane glycoprotein (Extended Figure 4E). Higher annual mean concentrations of black carbon were associated with lower levels of IgGFc-binding protein (Beta: -0.18, 95% CI: -0.29, -0.06) (Extended Figure 4H).

Third, we performed another post-hoc sensitivity analysis that stratified the association between videogame playing and serpin family G member 1 by the season when participants responded to the questionnaire (April-October vs. November-March). The association remained significant in both periods (Supplemental Table 7). When participants responded to the questionnaire in April-October, participants who played videogames daily had higher levels of serpin family G member 1 compared to those who played a few times or less (Beta: 0.13, 95% CI: 0.02, 0.25). When participants responded to the questionnaire in November-March, participants who played videogames a few times a month (Beta: 0.14, 95% CI: 0.03, 0.25) and daily (Beta: 0.15, 95% CI: 0.05, 0.26) also had higher levels of serpin family G member 1. By the Chow test, there was no significant difference between the coefficients in the two groups. After further adding interaction terms between videogame playing and time period to the model, the interaction terms were not significant.
