## Supplementary figures and images for "Associations between the mid-adolescent external exposome and proteomic biomarkers of mental health"

### Extended Figure 1.tiff

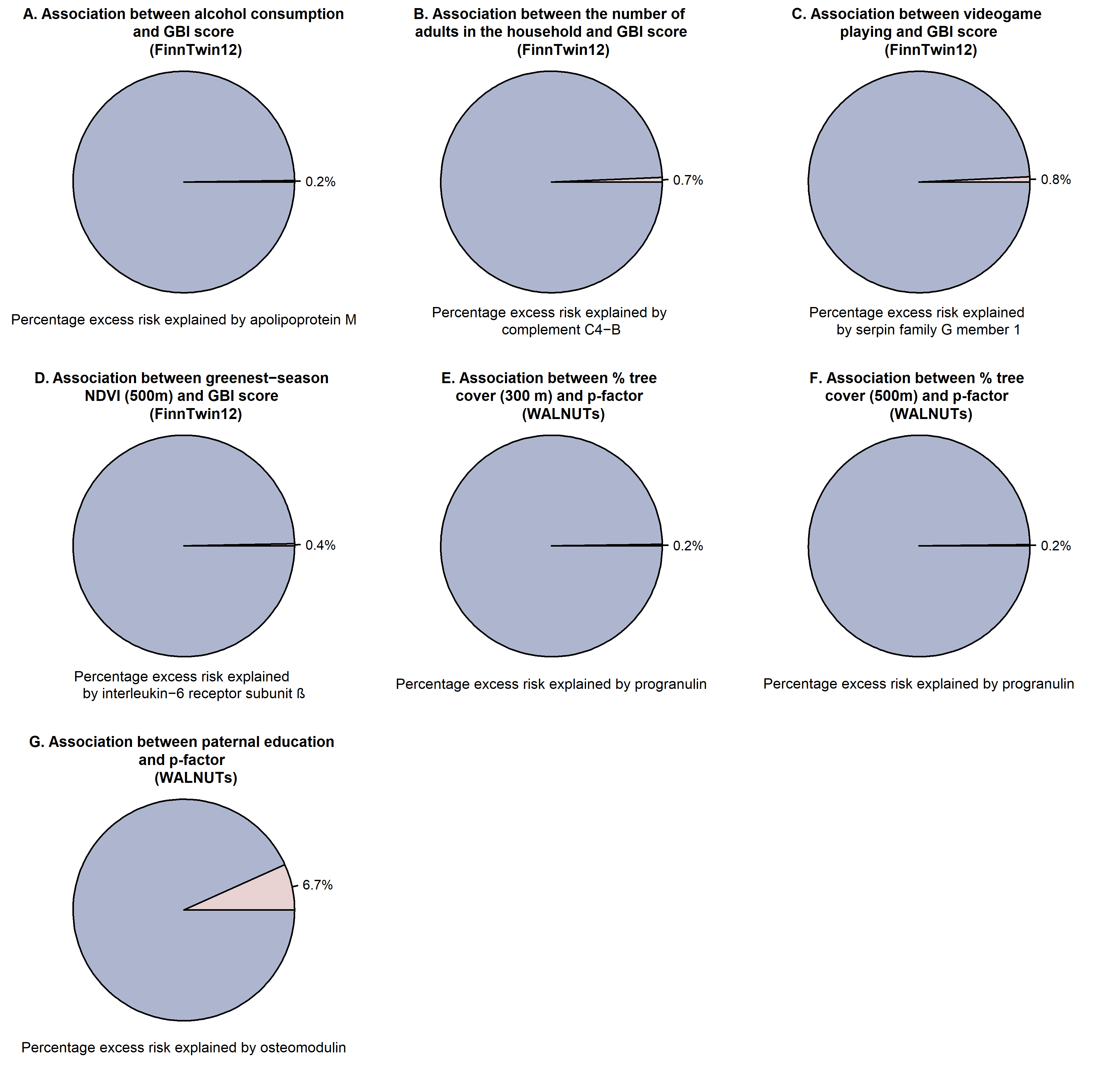

### Extended Figure 2.tiff

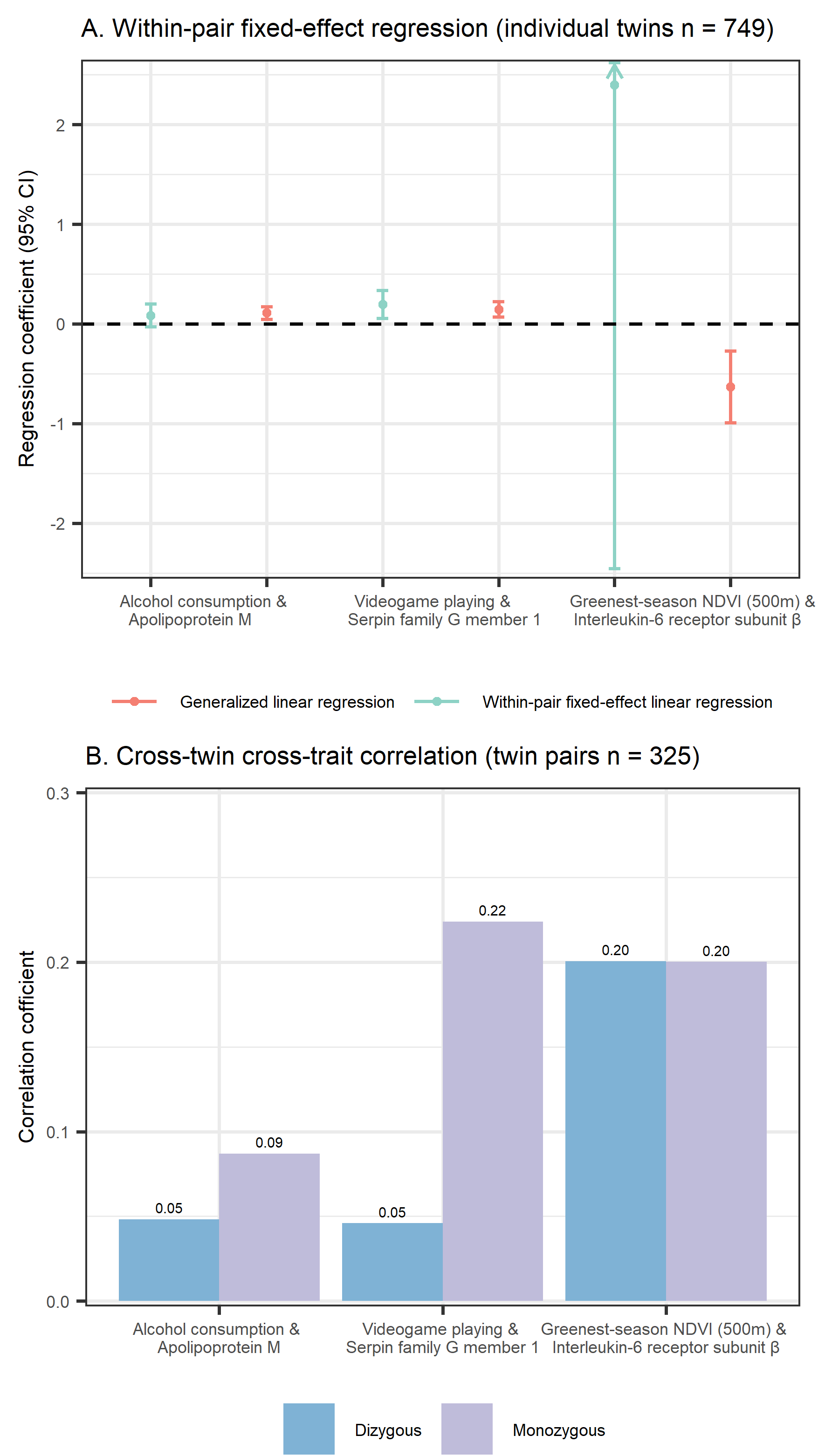

### Extended Figure 3.tiff

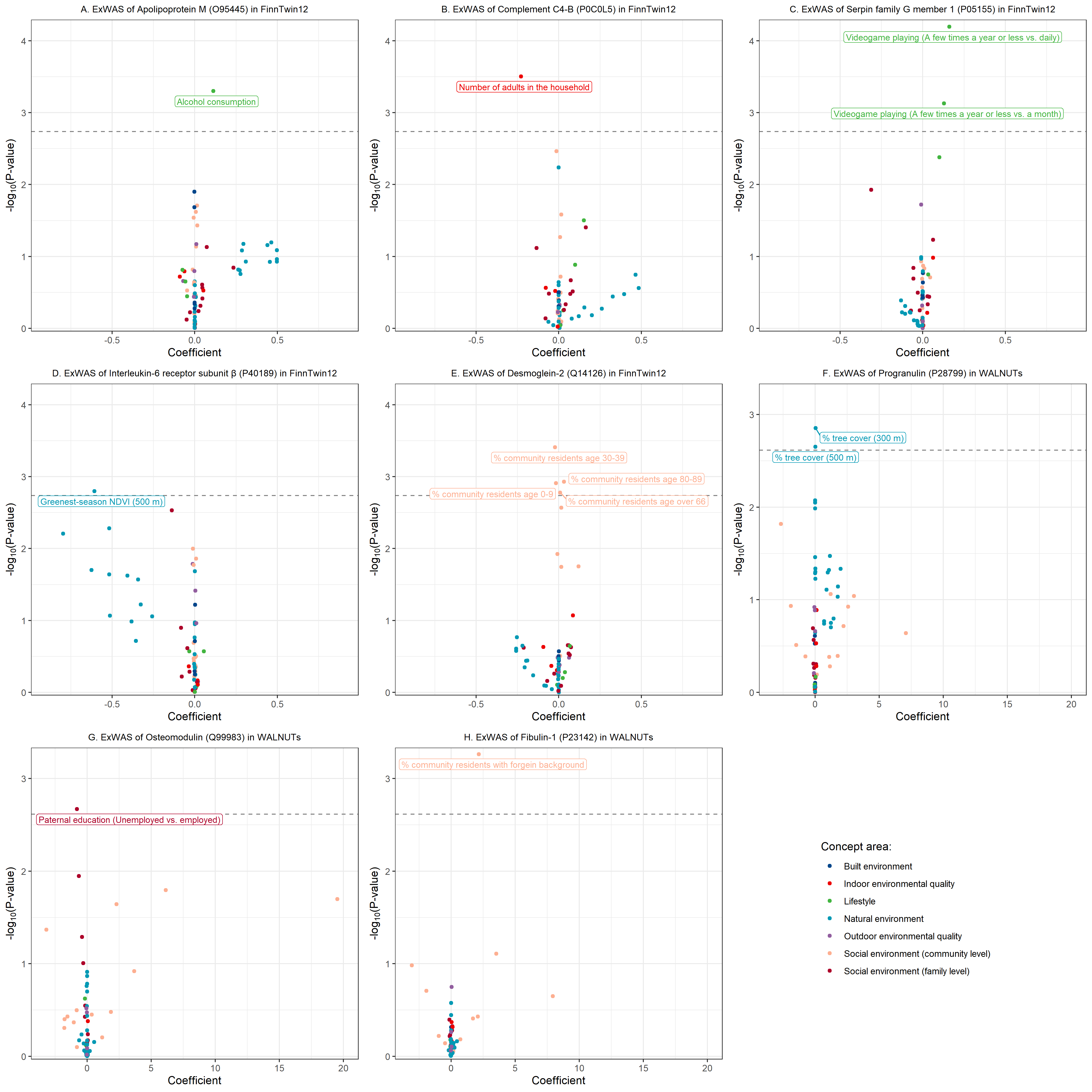

### Extended Figure 4.tiff

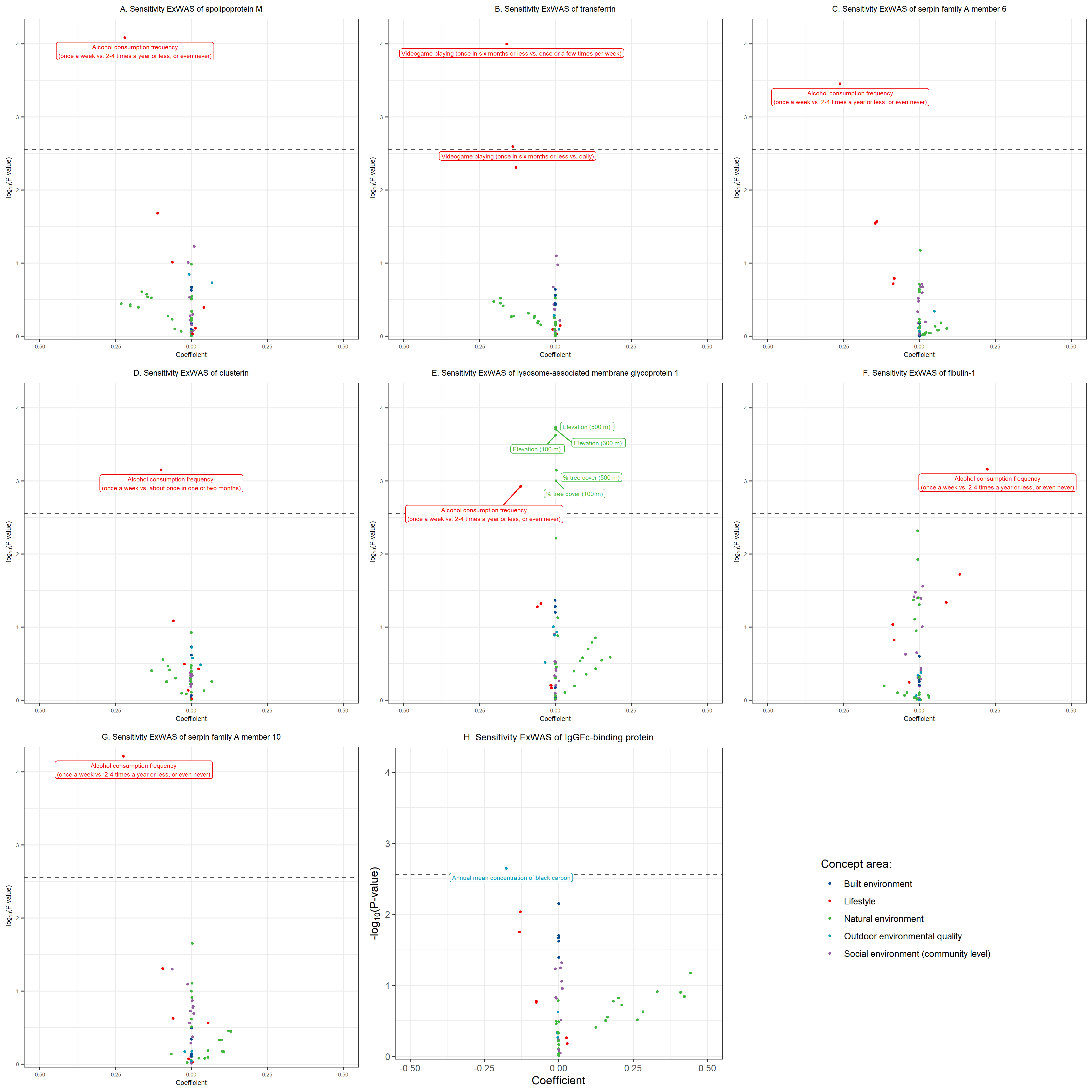
